# Diagnostic accuracy of the STANDING algorithm in patients with isolated vertigo/dizziness, a multicentre prospective study (STANDING-M)

**DOI:** 10.1101/2024.12.21.24318888

**Authors:** Mattia Ronchetti, Paola Bartalucci, Giuseppe Pepe, Giulia Canaroli, Simone Magazzini, Ersilia de Curtis, Federico Di Sacco, Maurizio Bartolucci, Rudi Pecci, Claudia Casula, Lorenzo Pelagatti, Ginevra Fabiani, Andrea Pavellini, Cosimo Caviglioli, Peiman Nazerian, Paolo Vannucchi, Simone Vanni

## Abstract

**Aim:** To evaluate the diagnostic accuracy of the STANDING algorithm across different emergency departments (ED)s. As secondary outcomes we compared the STANDING and the local usual care (LUC), in term of accuracy, use of diagnostic resources and length of stay (LOS).

**Methods:** We prospectively enrolled adult patients presenting with vertigo/dizziness at one ‘hub’ and three ‘spoke’ EDs in Tuscany, evaluated using either STANDING or LUC depending on the availability of a trained emergency physician (EP). Imaging tests, consultations and discharge/admission decisions were made independently of the study. The reference standard was a diffusion-weighted MRI of the brain and 30-days follow-up.

**Results:** We included 456 patients, 242 (53%) assessed by STANDING. No difference in age, gender and prevalence of cardiovascular risk factors were present between STANDING and LUC groups. The prevalence of central vertigo was 8.6%, with ischemic stroke (4.2%) as the leading cause, without differences between the two groups. The sensitivity, specificity, positive and negative predictive values (95% CI) of STANDING for central disease were 88.2% (63.6-98.5), 91.6% (87.1-94.8), 44.1% (33.2-55.7), 99% (96.5-99.7), without differences between the ‘hub’ and the ‘spoke’ centres and when only ischemic stroke was considered. STANDING demonstrated higher specificity and positive predictive values than that of LUC (36.5% and 14.7%, p<0.05 for both). Additionally, requests for head CT were lower (48.3% vs. 66.8%) and LOS shorter (289 vs. 351 minutes) in the STANDING group (p<0.05 for both).

**Conclusions:** The STANDING algorithm showed a good accuracy and a very high negative predictive value for excluding central disease and stroke, across different EDs. Compared to LUC, STANDING showed increased specificity, reduced utilisation of head CT and a shorter LOS.

## INTRODUCTION

Vertigo is defined as an illusory sensation of movement of oneself or one’s surroundings when no actual movement is occurring [1]. Dizziness refers to the sensation of disturbed or impaired spatial orientation without a false or distorted sense of motion [1]. These terms are often used interchangeably in clinical practice and they are common reasons for medical consultation, accounting for up to 3% of emergency department (ED) visits [2]. The majority of these cases are attributed to benign conditions such as benign paroxysmal positional vertigo (BPPV), acute unilateral peripheral vestibulopathy (AUPV), and Menière’s disease [3-5]. However, a significant minority of patients, ranging from 2% to 6%, are diagnosed with central nervous system pathologies, such as ischemic stroke [6-8]. Among ED patients, vertigo and dizziness are the symptoms most commonly associated with missed stroke diagnoses and delays in diagnosis can significantly increase morbidity and mortality [9,10].

Some diagnostic algorithms have been proposed for evaluating patients with acute vertigo or dizziness, including the STANDING algorithm [11-14]. This algorithm assesses a broad range of patients presenting with vertigo/dizziness, in particular those with suspected BPPV (the most common form), acute vestibular syndrome (AVS), or imbalance without nystagmus through the evaluation of their standing position and gait. The STANDING algorithm has demonstrated high sensitivity and specificity in a monocentric study [14], and it has recently been externally validated by a French research group [15]. Other algorithms have also been proposed and externally validated, such as the HINTS [15, 21], and recent guidelines and consensus documents on this topic suggested to use them [5,6]. Despite these indications, many physicians in Italian and European EDs do not use validated diagnostic algorithms, questioning their applicability in daily clinical practice.

This multicenter study aimed to evaluate the diagnostic accuracy of the STANDING algorithm across different hospital settings and to compare the diagnostic pathway and 30-day outcomes of patients assessed using the STANDING algorithm versus those evaluated through usual clinical practice.

## MATERIALS AND METHODS

### Study design and setting

The study, registered on ClinicalTrials.gov (registration number NCT06515951), is a prospective, multicenter, non-profit investigation aimed at evaluating the diagnostic accuracy of the STANDING algorithm in patients presenting with vertigo or instability. Patient recruitment occurred between June 2022 and June 2023 across four ED in the Tuscany region of Italy. These included a teaching hospital and hub center for stroke and trauma, Careggi University Hospital in Florence, which manages approximately 110,000 presentations annually, as well as three community hospitals—Santo Stefano Hospital (Prato), San Giuseppe Hospital (Empoli), and Versilia Hospital (Viareggio)—which collectively handled around 220,000 presentations. The study population did not overlap with those from previous clinical studies. The study protocol received approval from the local ethics committee (N°ID 21817) and all participants provided informed consent in accordance with the Declaration of Helsinki.

The study focused on adult patients presenting to the ED with acute vertigo or dizziness, defined according to the Bárány Society [1], with an onset within one week.

Exclusion criteria included: 1) symptoms that could be immediately attributed to a non-neuro-otological pathology (e.g., anemia, metabolic disorders); 2) conditions affecting the cervical spine or neck that contraindicated diagnostic maneuvers; 3) age under 18 years; 4) a life expectancy of less than 3 months or an inability to complete a 30-day follow-up; 5) the presence of clinically overt neurological signs identified during triage; 6) asymptomatic presentation at the time of the visit; 7) inability to participate in the study (e.g., severe dementia) and 8) refusal to participate.

### Primary and secondary Outcomes

The primary outcome of the study was to evaluate the diagnostic accuracy—in particular, the sensitivity and specificity—of the STANDING algorithm when performed by trained emergency physicians (EPs) to diagnose a central cause of isolated vertigo or dizziness.

Planning to enrol consecutive patients with vertigo/dizziness who were not evaluated by STANDING due to the absence of a trained EP on duty (local usual care, LUC), the secondary outcomes of the present study were as follows:

- Comparison of diagnostic accuracy: The difference in diagnostic accuracy between the STANDING cohort and the LUC cohort.
- Use of diagnostic resources and Length of Stay (LOS): the comparison between the STANDING and LUC in terms of the number of brain imaging tests performed, the number of Ear-Nose-Throat (ENT) or neurological consultations requested, and the LOS in the ED.
- Adverse events during 30-Day follow-up of patients discharged from the ED with a diagnosis of “benign” vertigo, including all-cause mortality, diagnoses of central vertigo or stroke, reperfusion therapy of the brain vessels (thrombolysis or catheter directed therapy), neurosurgery, and readmission for vertigo of suspected central origin within 30 days after ED discharge.

Inclusion and exclusion criteria were initially assessed by a triage nurse, who then assigned the patient to a physician. Depending on the availability of a physician trained in the STANDING protocol in that moment in ED, it was applied the STANDING protocol (STANDING cohort) otherwise the patient followed a usual clinical practice (LUC). To maintain a balanced study design, efforts were made to achieve a 1:1 ratio between the two cohorts by using an anonymized list to assign patients.

At least three emergency physicians per center were trained in the STANDING algorithm before the start of the study. The training program included a six-hour workshop consisting of four hours of theoretical instruction and two hours of practical sessions with healthy volunteers. This training was supplemented by ten supervised examinations in the ED, conducted under the guidance of a vestibologist or an experienced EP.

In both the STANDING and LUC cohorts, the attending EP determined the necessity of blood tests, diagnostic imaging, specialist consultations, therapies, hospitalization or patient discharge. These decisions were made independently of the patient’s participation in the study. Discharged patients were instructed to return to the hospital if their symptoms changed, worsened, or reappeared.

All patients included in the study were re-evaluated at 30 days by one of the expert team. A clinical evaluation was preferred whenever possible. If direct examination was not feasible, patients were contacted by telephone to assess the persistence of vertigo and the occurrence of adverse events, including death from any cause, diagnosis of central vertigo or stroke, the need for thrombolysis or mechanical thrombectomy, neurosurgery, or ED readmission for vertigo/dizziness resulting in hospitalization. If a patient could not be reached, their electronic medical records were reviewed to check for any ED re-access or hospitalizations related to stroke or other brain pathologies, with specific attention to 1) stroke diagnosis, 2) revascularization procedures such as thrombolysis or thrombectomy, and 3) any neurosurgical interventions. Patients for whom no data could be obtained were considered lost to follow-up.

### The STANDING algorithm

The STANDING algorithm is a diagnostic tool designed for evaluating vestibular disorders, utilizing clinical signs and bedside maneuvers in a four-step sequence:

1. **Assessment of Nystagmus: Identify the presence and type of nystagmus (spontaneous, positional, or absent).**
2. **Direction of Spontaneous Nystagmus: Determine the direction of the observed nystagmus.**
3. **Head Impulse Test: Evaluate the vestibulo-ocular reflex using the Head Impulse Test (HIT).**
4. **Assessment of Standing and Gait: Examine the patient’s standing and gait.**

**(1) Assessment of Nystagmus**: Nystagmus is evaluated using Frenzel lenses with the patient in the supine position. If spontaneous nystagmus is not detected, positional nystagmus is assessed first using the Pagnini-McClure test, followed by the Dix-Hallpike test. Positional paroxysmal nystagmus lasting 1-2 minutes with a beat direction aligned with the plane of the affected canal is indicative of benign paroxysmal positional vertigo.

**(2) Direction of Spontaneous Nystagmus**: If spontaneous nystagmus is present, its direction is assessed. Multidirectional or vertical nystagmus suggests central vertigo.

**(3) Head Impulse Test (HIT)**: Only when spontaneous nystagmus is horizontal and unidirectional, the HIT is applied. In the presence of an acute unilateral labyrinthine lesion, a corrective eye movement (saccade) towards the reference point will be observed following a rapid head turn in the affected direction. A positive HIT points to a peripheral disorder, while a negative HIT suggests central vertigo.

**(4) Assessment of Standing and Gait**: Following the nystagmus assessment, all patients and especially those without detectable nystagmus are asked to stand in order to evaluated their gait. Objective inability to sit, stand or walk without support or falling raises suspicion of central nervous system pathology [19].

The STANDING protocol is considered indicative of central vertigo ("worrisome STANDING") when at least one of the following criteria is met: (1) spontaneous vertical or multidirectional nystagmus; (2) spontaneous horizontal unidirectional nystagmus with a negative HIT; or (3) inability to sit, stand or walk without support, but also those in class 1 of Carmona (mild to moderate imbalance with walking independently) in the absence of detectable nystagmus. Patients in the LUC cohort are classified as having either “benign” or “worrisome” results based on the local clinical usual evaluation according to the attending EP’s usual practice.

### Reference standard for the diagnosis of central vertigo

The reference standard for diagnosing central vertigo was magnetic resonance imaging (MRI) of the brain and brainstem with diffusion-weighted imaging (DWI). For patients with contraindications to MRI, computed tomography (CT) with contrast and, if necessary, angiographic study (CTA) was used. If contrast medium was contraindicated due to severe allergy or renal failure, CT without contrast (NCCT) was performed. Central vertigo diagnosis was based on the identification of an acute brain lesion via neuroimaging. Acute ischemic stroke was diagnosed based on restricted diffusion on DWI-MRI or a hypodense area with ischemic damage characteristics on CT, congruent with the patient’s symptoms.

Patients discharged directly from the ED were instructed to undergo DWI-MRI of the head within 10 days. If DWI-MRI was not performed, the final diagnosis of central vertigo was determined by an expert panel, including an emergency physician, a radiologist and a neurologist, based on clinical and instrumental data, as well as any developments during the one-month follow-up and the results of the follow-up visit. In cases where a patient died before undergoing neuroimaging, they were classified as having central vertigo.

### Sample size

Based on data from the previous monocentric study [14], the estimated prevalence of central pathology in the sample is approximately 10%. To ensure the inclusion of about 20 patients with central pathology per group and to achieve 95% confidence intervals for sensitivity or specificity estimates within 10%, with an anticipated 10% follow-up dropout rate, it was necessary to enroll 220 patients per group. Considering the annual patient census of approximately 330,000 across four centers, with 1% presenting vertigo/dizziness (3,300) and accounting for exclusions (15% due to criteria 1) to 5), 30% due to non-vestibular causes and 20% due to the unavailability of EPs participating in the study, it was projected that centers would enroll 440 patients and complete data collection within 12 months.

### Statistical plan

Continuous variables were expressed as mean ± standard deviation (SD), and dichotomous variables as percentages with 95% confidence intervals. The diagnostic accuracy of the STANDING test for central vertigo was assessed by calculating sensitivity, specificity, positive and negative predictive values, and positive and negative likelihood ratios, each with 95% confidence intervals. Secondary outcomes included the percentages of brain imaging tests, neurological or ENT consultations requested in the ED, the LOS in the ED excluded the time in the observation unit and combined adverse events among patients discharged from the ED. Patients lost to follow-up were excluded from the statistical analysis. Comparisons of dichotomous variables were performed using χ2
 or Fisher’s exact test for values <5%. Comparisons of continuous variables were conducted using Student’s t-test or Mann-Whitney test for non-normally distributed data, with p < 0.05 considered statistically significant. Calculations were performed using SPSS software (version 22.0, SPSS, Chicago, Illinois, USA).

## RESULTS

Among the 1,098 patients screened for inclusion, 629 (57%) were excluded based on the exclusion criteria and 469 (43%) were eligible for the study (Figure 1). Of these, 13 patients (2.7%) were lost to follow-up. Consequently, 456 patients were included: 242 (53%) were evaluated by EPs trained in the STANDING algorithm, while 214 (47%) received LUC (Figure 1).

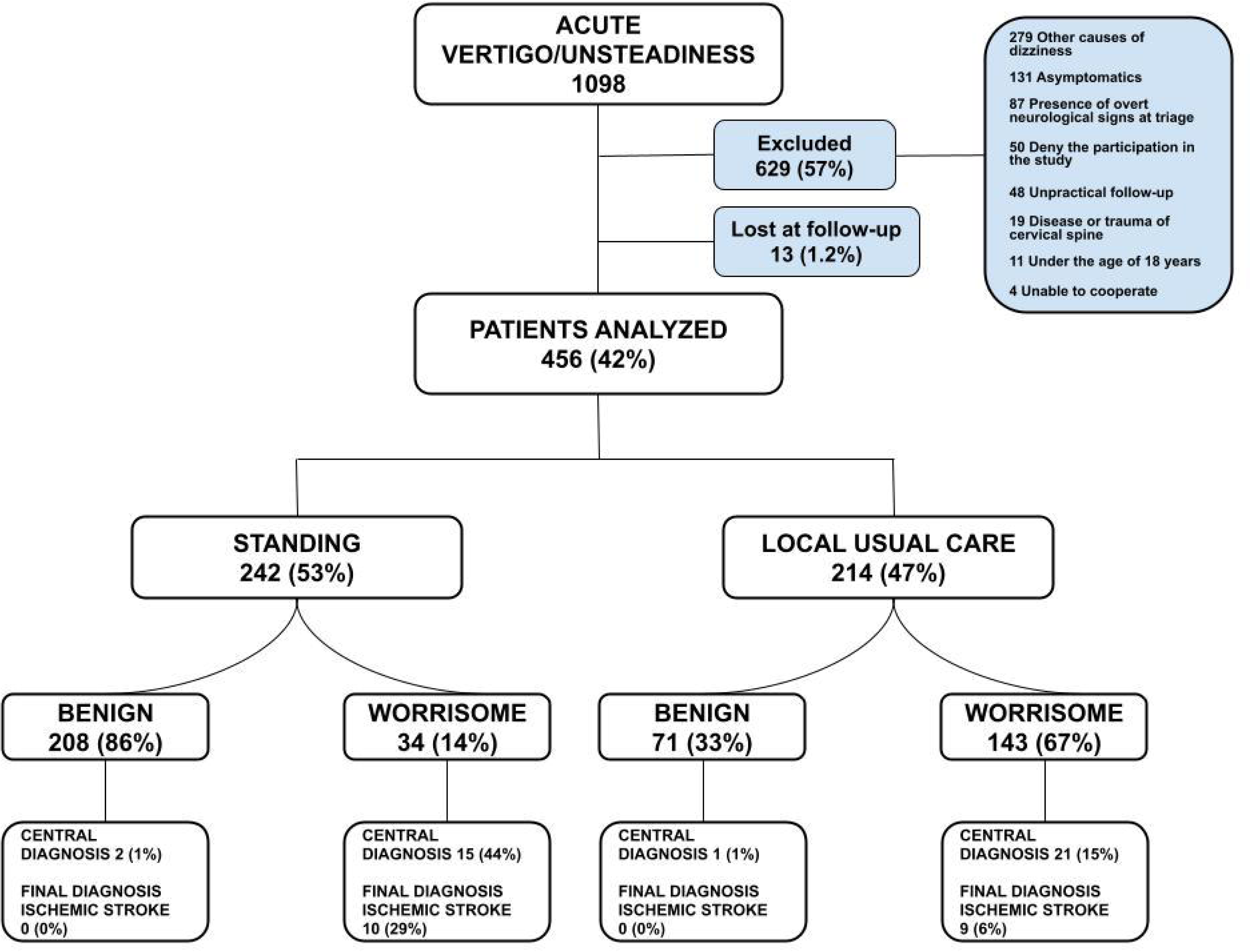

### General Characteristics of patients investigated

The prevalence of cardiovascular risk factors was similar between the two groups, with atrial fibrillation prevalence not differing significantly (Table 1). Regarding presenting symptoms, continuous vertigo (lasting hours to days) and headache were more frequently reported in the LUC cohort compared to the STANDING cohort (35% vs. 25.2%, p = 0.024; 19.6% vs. 8.7%, p < 0.01, respectively) (Table 1). The prevalence of subtle neurological symptoms was comparable between the groups. However, positional or spontaneous nystagmus was more commonly observed in patients evaluated by the STANDING algorithm compared to those in the LUC cohort (49.2% vs. 5.6%, p < 0.01; 21.5% vs. 12.1%, p < 0.01, respectively) (Table 1). Among the 242 patients evaluated by the STANDING algorithm, 208 (86%) had a benign profile, while 34 (14%) had a worrisome profile (Figure 1). In contrast, in the LUC cohort, 71 (33%) had a benign profile and 143 (67%) had a worrisome profile (p < 0.01 vs. STANDING cohort).

**Table 1.** Clinical characteristics at presentation of included patients.

|  | **Overall**<br>**N = 456** | **STANDING**<br>**N = 242** | **LUC**<br>**N = 214** | **p-value** |
| --- | --- | --- | --- | --- |
| **Cardiovascular risk factors** |  |  |  |  |
| Hypertension | 186 (40.8%) | 98 (40.5%) | 88 (41.1%) | 0.924 |
| Diabetes | 33 (7.2%) | 17 (7.0%) | 16 (7.5%) | 0.859 |
| Atrial fibrillation | 33 (7.2%) | 13 (5.4%) | 20 (9.3%) | 0.107 |
| Previous TIA/stroke | 27 (5.9%) | 17 (7.0%) | 10 (4.7%) | 0.440 |
| Current cigarette smoking | 27 (5.9%) | 16 (6.6%) | 11 (5.1%) | 0.556 |
| Dyslipidemia | 76 (16.7%) | 46 (19.0%) | 30 (14.0%) | 0.167 |
| At least 1 CV risk factor | 237 (52.0%) | 127 (52.5%) | 110 (51.4%) | 0.851 |
| **Symptoms of presentation** |  |  |  |  |
| Continuous vertigo | 136 (29.8%) | 61 (25.2%) | 75 (35.0%) | 0.024 |
| Headache | 63 (13.8%) | 21 (8.7%) | 42 (19.6%) | < 0.001 |
| Hearing loss | 28 (6.1%) | 20 (8.3%) | 8 (3.7%) | 0.051 |
| Vomiting | 212 (46.5%) | 118 (48.8%) | 94 (43.9%) | 0.347 |
| **Vital signs** |  |  |  |  |
| Heart rate (bpm ± SD) | 75 ± 13 | 74 ± 13 | 76 ± 12 | 0.110 |
| SBP (mmHg ± SD) | 142 ± 35 | 142 ± 21 | 142 ± 45 | 0.997 |
| DBP (mmHg ± SD) | 79 ± 11 | 80 ± 10 | 78 ± 11 | 0.213 |
| **Neurological signs** |  |  |  |  |
| Cranial nerve dysfunction | 19 (4.1%) | 10 (4.1%) | 9 (4.2%) | 1.000 |
| Limb weakness/hypoesthesia | 11 (2.4%) | 7 (2.9%) | 4 (1.9%) | 0.552 |
| Dysarthria/dysphagia | 5 (1.1%) | 4 (1.7%) | 1 (0.4%) | 0.377 |
| Limb ataxia | 10 (2.2%) | 5 (2.1%) | 5 (2.3%) | 1.000 |
| Dynamic/static ataxia | 54 (11.8%) | 32 (13.3%) | 22 (11.2%) | 0.384 |
| No neurological signs | 386 (84.6%) | 205 (84.7%) | 181 (84.5%) | 1.000 |
| **Nystagmus** |  |  |  |  |
| Positional nystagmus | 131 (28.7%) | 119 (49.2%) | 12 (5.6%) | < 0.001 |
| Spontaneous nystagmus | 79 (17.3%) | 53 (21.9%) | 26 (12.1%) | < 0.001 |
| No nystagmus | 246 (53.9%) | 70 (28.9%) | 176 (82.2%) | < 0.001 |

bpm=beats per minute; CV=Cardiovascular; IQR=Interquartile range; LUC=Local Usual Care; SBP=Systolic Blood Pressure; DBP=Diastolic Blood Pressure; SD= Standard deviation; TIA=Transient.

### Final diagnosis

During the initial evaluation in the ED, 260 patients (57%) underwent brain imaging: 260 with NCCT of the head, 50 with computed tomography angiography (CTA) of the head and neck, and 28 with DWI-MRI of the brain. At the 30-day follow-up, no patients had died, and an additional 82 patients underwent brain imaging, including 10 with NCCT and 72 with DWI-MRI. This brought the total number of patients evaluated by brain imaging to 287 (62.9%), with 160 (35%) assessed by the reference standard (DWI-MRI or CTA or NCCT in those with contraindications). For the remaining 296 patients (65%), the final diagnosis was determined by an expert panel (SV, MB, PB), based on clinical and instrumental data, and including any events occurring during the one-month follow-up (see above in the methods).

Among patients with central vertigo (39 cases, 8.6%), ischemic stroke was the most common diagnosis (19 cases; 48.7%), followed by neoplasms (6 cases, 15.4%) and demyelinating diseases (3 cases, 7.7%) (Table 2). Most ischemic strokes affected the cerebellum with the lateral medulla (14 cases; 73.7%) (Table 3) (Figure 2). For vertigo of other origins, BPPV was the most frequent pathology (258 cases; 56.6%), followed by AUPV in 14.3% of patients (Table 2).

**Table 2.**
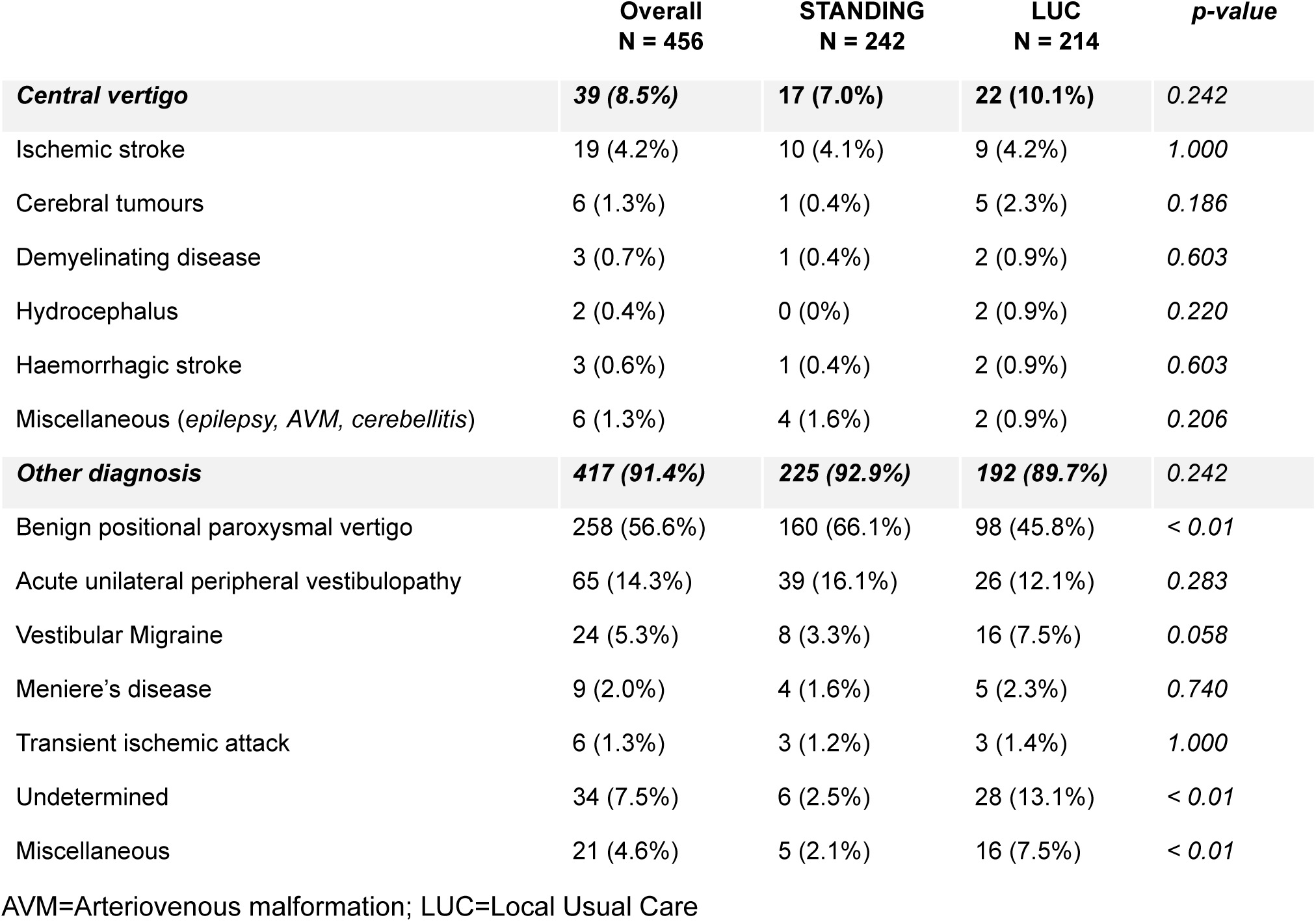
Main diagnosis of included patients.

**Table 3.** Features of patients affected by isohemic stroke.

| *Patient* | *Sex* | *Neurological signs at presentation* |  |  | *NIHSS* | *Ataxia* | *Nystagmus* | *Site* | *Etiology* |
| --- | --- | --- | --- | --- | --- | --- | --- | --- | --- |
| 1 | M | Left facial paralysis, hypoesthesia on the left side of the face, dysmetria, dysarthria. |  |  | 5 | 3 | Multidirectional nystagmus | Left inferior cerebellar hemisphere | Dissection |
| 2 | M | None |  |  | 0 | 1 | None | Lower right cerebellum | Cardioembolic |
| 3 | M | None |  |  | 0 | 3 | None | Left upper cerebellum | Cardioembolic |
| 4 | M | Facial paralysis, dysmetria |  |  | 3 | 2 | Gaze evoked nystagmus | Lower left cerebellum and posterior left bulb | Dissection |
| 5 | F | None |  |  | 0 | 1 | None | Lenticular and frontal subcortical capsule, insular region | Atherosclerotic |
| 6 | M | Mild facial paralysis |  |  | 1 | 1 | None | Right inferomedial cerebellar hemisphere, tonsil | Cardioembolic |
| 7 | M | None |  |  | 0 | 3 | None | Cerebellar vermis | Atherosclerotic |
| 8 | F | Diplopia |  |  | 0 | 1 | None | Upper cerebellum | Cardioembolic |
| 9 | F | None |  |  | 0 | 1 | Spontaneous horizontal and torsional nystagmus, HIT not done | Right inferior/ superior cerebellar hemisphere, tonsil, vermis. | Cardioembolic |
| 10 | M | Facial hypoesthesia |  |  | 1 | 1 | Multidirectional nystagmus | Pons | Dissection |
| 11 | F | None |  |  | 0 | 1 | Spontaneous horizontal nystagmus, HIT negative | Right cerebellar hemisphere and vermis | Atherosclerotic |
| 12 | M | None |  |  | 0 | 3 | None | Inferior cerebellar peduncle | Lacunar |
| 13 | M | None |  |  | 0 | 3 | None | Inferior cerebellar peduncle | Lacunar |
| 14 | F | None |  |  | 0 | 2 | None | Left cerebellar hemisphere | Cardioembolic |
| 15 | M | Right facial deficit, right upper limb weakness |  |  | 2 | 3 | None | None (treated by rTPA) | Atherosclerotic |
| 16 | F | Facial paralysis, dysmetria, dysarthria |  |  | 3 | 3 | Multidirectional nystagmus | Right inferior cerebellar hemisphere | Atherosclerotic |
| 17 | F | None |  |  | 0 | 2 | None | Right superior cerebellar peduncle | Cardioembolic |
| 18 | M | None |  |  | 0 | 2 | Spontaneous horizontal nystagmus, HIT negative | Vermal and right paravermal region | Dissection |
| 19 | M | None |  |  | 0 | 2 | Spontaneous horizontal nystagmus, HIT not done | Left upper cerebellar hemisphere | Dissection |

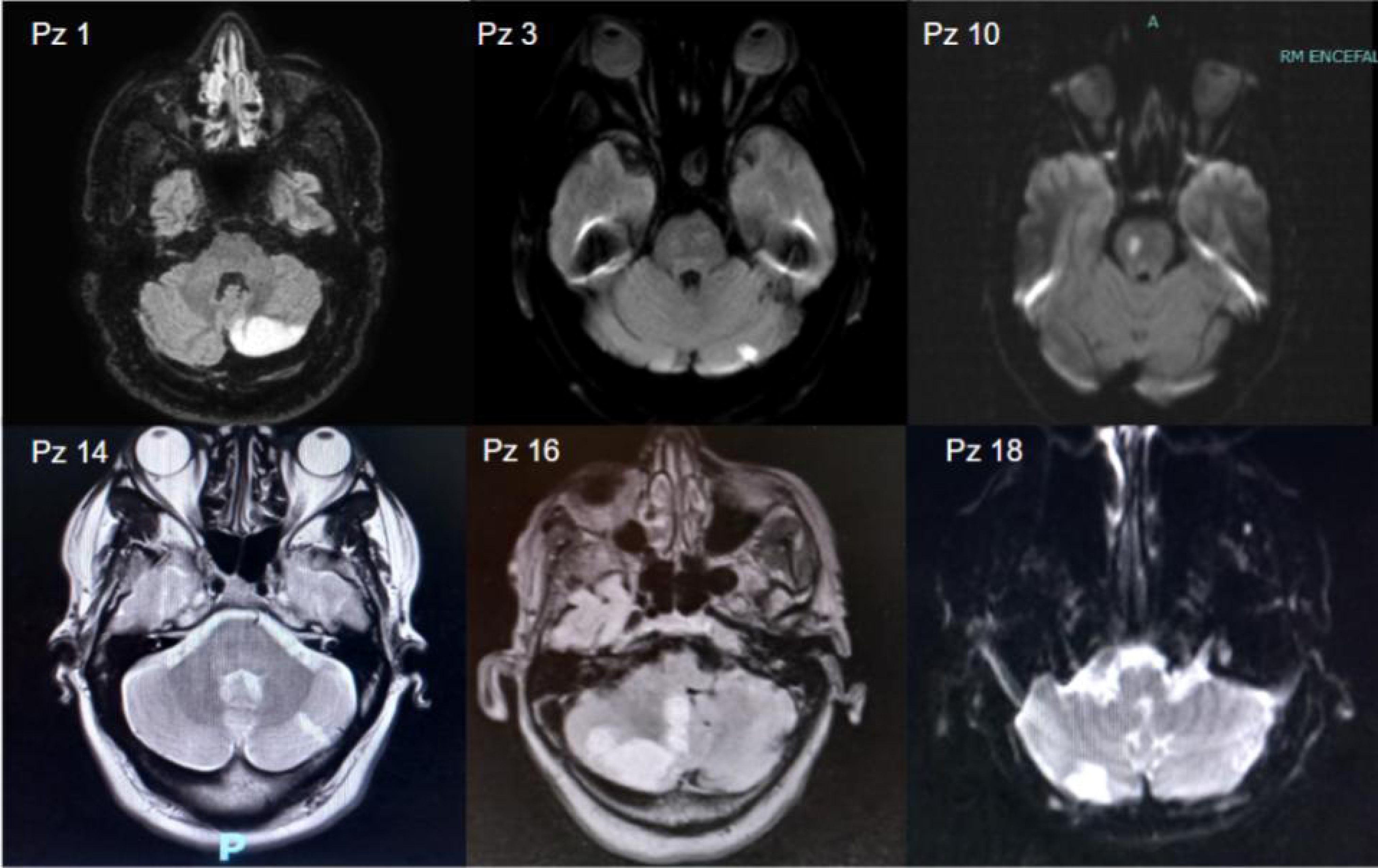

Among the patients affected by a central vertigo, 31 (79.5%) had a National Institutes of Health Stroke Scale (NIHSS) score of 0. Only 12 patients (30.7%) exhibited at least one neurological sign as detailed in Table 1, 14 patients (35.9%) if we include a grade 2-3 ataxia within the neurological signs, and 28 patients if we include a grade 1 ataxia without spontaneous or positional nystagmus (71.7%). Thus, 28.3% of patients with a final diagnosis of central vertigo showed at first evaluation a normal standard neurological examination if we don’t consider nystagmus.

### Accuracy of the STANDING algorithm

Among the patients with a "benign" profile only 2 (1%) were later diagnosed with central disease (Figure 1): one initially diagnosed with AUPV was subsequently identified with multiple sclerosis, and another initially diagnosed with BPPV was later found to have viral cerebellitis. Both patients underwent NCCT of the head in the ED, which did not reveal abnormalities. Among the 34 patients with a "worrisome" profile, 15 (44%) had central vertigo as defined by the presence of a lesion on brain imaging consistent with the patient’s symptoms, while 19 (56%) were false positives.

The STANDING algorithm achieved a sensitivity of 88.2%, specificity of 91.6%, positive predictive value (PPV) of 44.1%, and negative predictive value (NPV) of 99%, with similar performance for diagnosing ischemic stroke (Table 4). Diagnostic characteristics of STANDING did not differ in spoke centers (Table 4).

**Table 4.** Diagnostic accuracy of STANDING and comparison with Local Usual Care.

|  | **STANDING** | **STANDING spokes** | **LUC** |
| --- | --- | --- | --- |
| **All central causes (95% CI)** | **N=242** | **N=162** | **N=214** |
| *Sensitivity* | 88.2 (63.6-98.5) | 91.7 (62.9-99.6) | 95.5 (76.1-99.8) |
| *Specificity* | 91.6 (87.1-94.8) | 86.8% (81.7-89.3) | 36.5 (34.72-37.4)* |
| *PPV* | 44.1 (33.2-55.7) | 50% (34.3-54.3) | 14.7 (11.7-15.3)* |
| *NPV* | 99.0 (96.5-99.7) | 98.6% (93.6-99.9) | 98.6 (92.6-99.9) |
| *LR+* | 10.4 (6.6-16.6) | 6.6 (3.4-7.8) | 1.5 (1.2-1.6)* |
| *LR-* | 0.1 (0.0-0.5) | 0.1 (0.0-0.5) | 0.1 (0.1-0.7) |
| **Ischemic Stroke (95% CI)** |  |  |  |
| *Sensitivity* | 100 (69.1-100) | 100 (65.6-100) | 100 (63.7-100) |
| *Specificity* | 89.7 (85.0-93.3) | 84.1 (80.4-84.1) | 34.6 (33-34.6)* |
| *PPV* | 29.4 (22.2-37.8) | 40.9 (26.8-40.9) | 6.3 (4-6.3)* |
| *NPV* | 100 (98.2-100) | 100 (95.5-100) | 100 (94.9-100) |
| *LR+* | 9.7 (6.6-14.1) | 6.3 (3.3-6.3) | 1.5 (0.9-1.5)* |
| *LR-* | 0 (0-0.4) | 0 (0-0.5) | 0 (0.0-1.1) |

\*= p<0.05. PPV=Positive predictive value; NPV=Negative predictive value; LR=Likelihood ratio; LUC=Local Usual Care.

In comparison, only 33% of patients evaluated according to LUC were classified as “benign”, whereas the majority of patients were considered as “worrisome” (p<0.01 vs STANDING cohort). Looking at diagnostic accuracy, LUC correctly identified 21 out of 22 cases of central vertigo, but only 70 out of 192 cases with other diagnoses, showing similar sensitivity (95.5%) but lower specificity (36.5%) compared to the STANDING cohort (Table 4). The PPV and positive likelihood ratio (PLR) for LUC were also significantly lower than those for the STANDING algorithm (Table 3). Additionally, LUC was associated with a significantly higher number of undetermined cases (13.1% vs 2.5%)(Table 2).

### Analysis of secondary outcomes

Head CT was performed less frequently in the STANDING cohort compared to the LUC cohort (48.3% vs. 66.8%; p < 0.01), while the proportion of patients undergoing urgent DWI-MRI was slightly higher in the STANDING cohort (Table 5).

**Table 5.** Secondary outcomes and of the length of stay (LOS) in ED.

|  | Overall<br>N = 456 | STANDING<br>N = 242 | LUC<br>N = 214 | p-value |
| --- | --- | --- | --- | --- |
| **Initial imaging tests** |  |  |  |  |
| Head NCCT | 260 (57.0%) | 117 (48.3%) | 143 (66.8%) | $< 0.01$ |
| Head CTA | 50 (10.9%) | 26 (10.7%) | 24 (11.2%) | 0.872 |
| Head DW-MRI | 28 (6.1%) | 20 (8.3%) | 8 (3.7%) | 0.050 |
| **Specialist consultations** |  |  |  |  |
| ENT or vestibologist | 63 (13.8%) | 28 (11.6%) | 35 (16.4%) | 0.174 |
| Neurology | 50 (11.0%) | 23 (9.5%) | 27 (12.6%) | 0.298 |
| Overall consultation | 113 (24.8%) | 51 (21.1%) | 62 (29.0%) | 0.068 |
| **Length of stay** |  |  |  |  |
| Mean±SD (minutes) | 318±221 | 289±202 | 351±236 | $< 0.01$ |

CTA=Computed tomography angiography; DW-MRI=Diffusion-weighted magnetic resonance imaging; ENT=ear, nose, and throat doctor; SD=Standard Deviation; LUC=Local Usual Care; NCCT= Non Contrast Computer tomography.

Specialist consultations were requested in 24.8% of cases, with fewer requests in the STANDING cohort (21.1%) compared to the LUC cohort (29%) (Table 5). LOS in the ED was significantly shorter for patients evaluated using the STANDING algorithm (290 minutes vs. 348 minutes; p<0.05) (Table 5).

Among patients discharged with a benign profile in the STANDING cohort, there were no deaths, strokes, or instances of reperfusion or neurosurgical treatments during follow-up. Four patients (2%) were hospitalised during follow-up including two for the persistence of symptoms caused by BPPV, one for viral cerebellitis and one for endocarditis (Table 6).

**Table 6.**
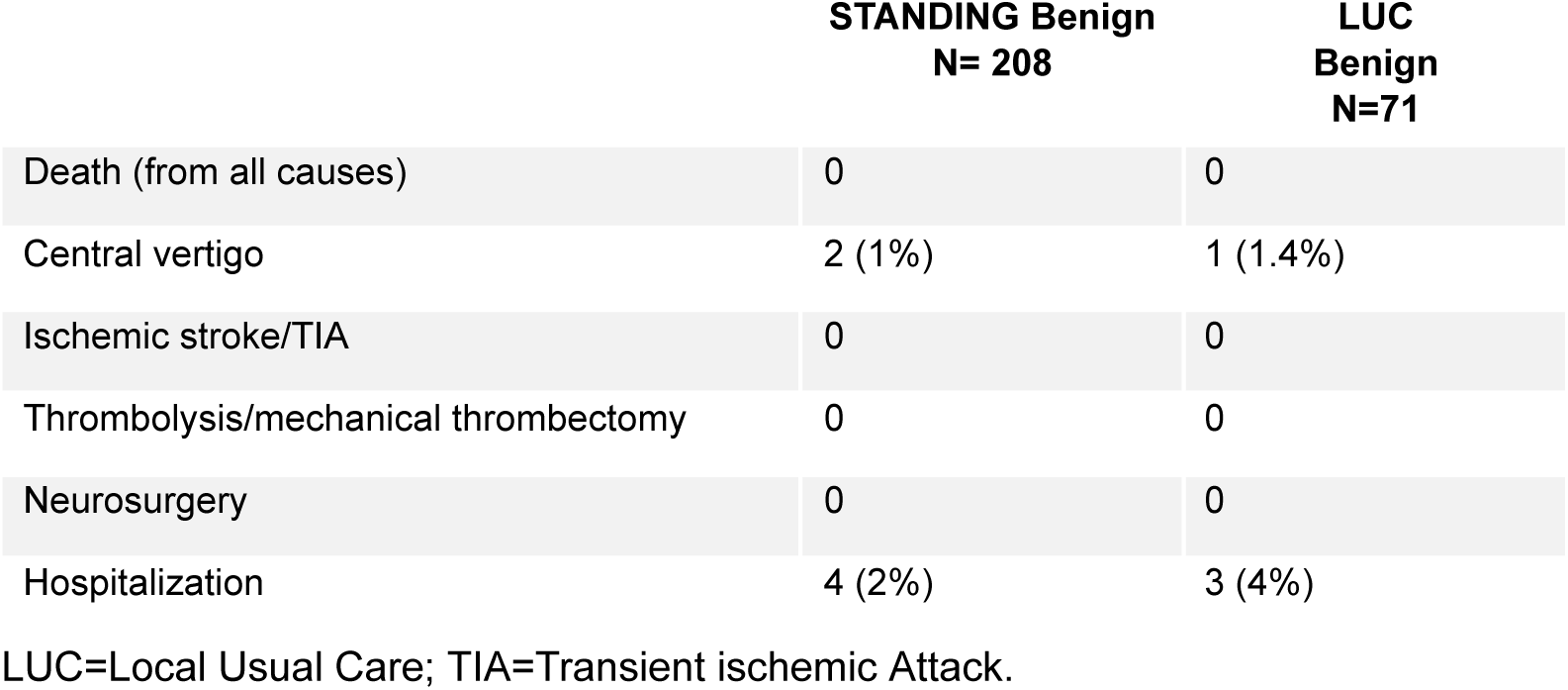
Adverse events during 30-days follow-up in patients discharged from ED with a benign profile.

## DISCUSSION

This multicenter study demonstrated that the STANDING algorithm accurately identifies patients with isolated vertigo or dizziness who require urgent evaluation for potential central diseases, particularly stroke, across different types of ED (Spokes vs Hub). Moreover, this study builds on previous research by extending the known diagnostic value of the STANDING algorithm, suggesting that the algorithm exhibited similar sensitivity but higher specificity compared to usual clinical care.

For the first time, STANDING has been evaluated in a multicenter setting, including community, non-teaching hospitals, confirming its accuracy when used by properly trained emergency physicians. This aligns with findings from a previous monocentric validation study [14] and an external validation study conducted in France [15]. In this multicenter study, STANDING’s sensitivity (88%) was slightly lower than that reported in a recent meta-analysis of neurological examinations for vertigo or dizziness (93-100%), but specificity remained consistent [18]. Similarly, Gerlier et al. [19] reported even lower sensitivity (84%) when STANDING was performed by interns, attributing this to varying levels of training and confidence. The modest reduction in sensitivity observed in our study may reflect the relative inexperience of the physicians trained for this study and differences in training compared to the previous study [14]. The training for new emergency physicians in this multicenter study consisted of a six-hour workshop with 10 proctored examinations, omitting the one-month supervised use under an expert physician, as was done in the monocentric study [14]. The shorter training could have reduced the investigators’ performance. The potential dependence of algorithm performance on the number of patients evaluated per month by each operator was recently highlighted in another paper from our group [20].

Despite the slightly lower sensitivity for central diseases, the specific sensitivity for stroke remained high (over 90%) even in this study and more than two-thirds of the patients were enrolled in community hospitals.

By aggregating data from different centers, this study allowed for a precise estimate of the prevalence of the most frequent diseases presenting with isolated vertigo or dizziness in the ED. We found that BPPV was the most common diagnosis, accounting for over 50% of patients, followed by AUPV (14%) and migraine (4.8%), collectively representing three-quarters of the selected population. The fact that more than 50% of patients with isolated vertigo or dizziness in the ED are affected by BPPV of the posterior (31.8%) or lateral (24.8%) canals underscores the utility of training emergency physicians in the clinical maneuvers necessary for diagnosing these conditions. Diagnostic algorithms for these patients should include such maneuvers. The fourth most common diagnosis, and the first among central causes, was ischemic stroke, with a prevalence of 4.2% (95% CI: 2.5%-6.4%). This prevalence is comparable to our previous studies [13,14] and aligns with recent meta-analysis findings (3.2%-6%) [18]. The stroke incidence we observed is lower than that reported by Gerlier et al. (16.3%) [15]. This discrepancy could be due to the inclusion of a population with higher cardiovascular risk in the French study, as indicated by a higher proportion of patients with previous stroke or TIA (13.6% vs 5.9%) and those with more than one cardiovascular risk factor (29.2% vs 13.6%).

Unlike previous prospective studies [14,15], our investigation included patients who were not assessed using the STANDING algorithm, thereby facilitating comparisons with the usual care group. The integration of the STANDING algorithm into the diagnostic pathway was associated with an enhanced diagnostic yield, primarily attributable to its superior specificity compared to standard clinical practice. This improvement resulted in a higher rate of diagnoses of peripheral vestibular disorders, notably BPPV, which increased from 45.8% to 66.1%, alongside a decrease in undetermined cases (from 13.1% to 2.5%). We contend that the reduction in head NCCT scans and LOS can be largely ascribed to the increased specificity of the STANDING algorithm. This advancement is particularly noteworthy as STANDING incorporates diagnostic maneuvers for otolithic disorders and the evaluation of standing and gait setting it apart from other diagnostic algorithms such as HINTS or HINTS plus [11,21-22].

The study has some limitations. Firstly, this was not a clinical trial and patients were not randomized. Although the number of events and known risk factors appears balanced between the two groups (Table 1), we cannot exclude the possibility that other "unknown" variables may have biased our results, particularly regarding differences between the STANDING and LUC cohorts. Would a different study design have provided a higher level of evidence? In our opinion, even a pre-post observational study would not have guaranteed better evidence than our current design, due to potential changes in local diagnostic policies during the study period, especially in a multicenter study. Beyond the eventual planning of a randomized clinical trial, which is challenging for a clinical diagnostic algorithm for vertigo or dizziness, we believe that our study presents the best available evidence on the utility of systematic nystagmus evaluation performed by trained emergency physicians in ED. Another potential source of bias in comparing STANDING with usual care is that, according to the protocol, investigators applying the STANDING algorithm were free to manage patients independently of the algorithm’s results. This could have underestimated STANDING’s potential effect on the number of imaging tests and consultations, as evidenced by the number of NCCT scans performed in the ED in the STANDING cohort (48%) compared to the number of worrisome results (14%). Finally, similar to the STANDING validation study [14], not all patients included in this study underwent DWI-MRI, and about 65% of final diagnoses were based on expert consensus after 30-day follow-up. We acknowledge that this may reduce confidence in estimating the primary outcome (incidence of central vertigo or stroke). However, considering the very low rate of readmissions and the absence of new episodes of TIA or stroke during follow-up of patients with a "benign" STANDING diagnosis, we can reasonably exclude a significant loss of information.

In conclusion, STANDING proves to be an accurate method for evaluating acute isolated vertigo. The participation of diverse hospitals in this study enhances the algorithm’s applicability across various ED settings. A benign diagnosis made using STANDING significantly reduces the likelihood of serious conditions, like ischemic stroke, allowing for timely clinical evaluation and reduced use of head CT in the ED compared to LUC. These data support the wider implementation of validated algorithms in EDs and emphasize that accurate classification of vertigo and dizziness in the emergency setting relies mainly on clinical evaluation, including correct nystagmus assessment.

## Supporting information

Cover Letter

STARD Index

Study Protocol

## Data Availability

Dear Editor, all data analyzed during this study are included in the manuscript. Additional datasets supporting the findings of this study are available from the corresponding author upon reasonable request. Sincerely, Dr Mattia Ronchetti

## REFERENCES

1. Bisdorf A, Staab J, Newman-Toker D (2015) Overview of the international classification of vestibular disorders. Neurol Clin 33(3):541–550

2. Newman-Toker DE, Hsieh YH, Camargo CA Jr, Pelletier AJ, Butchy GT, Edlow JA (2008) Spectrum of dizziness visits to US emergency departments: cross-sectional analysis from a nationally representative sample. Mayo Clin Proc 83(7):765–775

3. Madlon-Kay DJ. Evaluation and outcome of the dizzy patient. J Fam Pract (1985) 21:109–13.

4. Herr RD, Zun L, Mathews JJ. A directed approach to the dizzy patient. Ann Emerg Med (1989) 18:664–72. 10.1016/S0196-0644(89)80524-4

5. Vanni, S., Vannucchi, P., Pecci, R. et al. Consensus paper on the management of acute isolated vertigo in the emergency department. Intern Emerg Med (2024). 10.1007/s11739-024-03664-x

6. Jonathan A Edlow, Christopher Carpenter, Murtaza Akhter et al. Guidelines for reasonable and appropriate care in the emergency department 3 (GRACE-3): Acute dizziness and vertigo in the emergency department

7. Lam JM, Siu WS, Lam TS, Cheung NK, Graham CA, Rainer TH. The epidemiology of patients with dizziness in an emergency department. Hong Kong J Emerg Med (2006) 13:133–9.

8. Kerber KA, Brown DL, Lisabeth LD, Smith MA, Morgenstern LB. Stroke among patients with dizziness, vertigo, and imbalance in the emergency department: a population-based study. Stroke (2006) 37:2484–7. 10.1161/01.STR.0000240329.48263.0d

9. Savitz SI, Caplan LR, Edlow JA. Pitfalls in the diagnosis of cerebellar infarction. Acad Emerg Med (2007) 14:63–8. 10.1111/j.1553-2712.2007.tb00373.x

10. Tarnutzer AA, Berkowitz AL, Robinson KA, Hsieh YH, Newman-Toker DE. Does my dizzy patient have a stroke? A systematic review of bedside diagnosis in acute vestibular syndrome. CMAJ (2011) 183:571–92. 10.1503/cmaj.100174

11. Kattah JC, Talkad AV, Wang DZ, Hsieh YH, Newman-Toker DE (2009) HINTS to diagnose stroke in the acute vestibular syndrome: three-step bedside oculomotor examination more sensitive than early MRI difusion-weighted imaging. Stroke 40(11):3504–3510

12. Newman-Toker DE, Edlow JA (2015) TiTrATE: a novel, evidence based approach to diagnosing acute dizziness and vertigo. Neurol. Clin. 10.1016/j.ncl.2015.04.011

13. Vanni S, Pecci R, Casati C, Moroni F, Risso M, Ottaviani M et al (2014) STANDING, a four-step bedside algorithm for differential diagnosis of acute vertigo in the emergency department. Acta Otorhinolaryngol Ital 34(6):419–426

14. Vanni S, Pecci R, Edlow JA, Nazerian P, Santimone R, Pepe G et al (2017) Differential diagnosis of vertigo in the emergency department: a prospective validation study of the STANDING algorithm. Front Neurol 8:590

15. Gerlier C, Fels A, Vitaux H, Mousset C, Perugini A, Chatellier G, Ganansia O (2023) Effectiveness and reliability of the four-step STANDING algorithm performed by interns and senior Eps for predicting central causes of vertigo. Acad Emerg Med 30(5):487–500

16. Sankalia D, Kothari S, Phalgune DS. Diagnosing stroke in acute vertigo: sensitivity and specificity of HINTS battery in Indian population. Neurol India. 2021;69:97–101.

17. Carmona S, Martinez C, Zalazar G, et al. The diagnostic accuracy of truncal ataxia and HINTS as cardinal signs for acute vestibular syndrome. Front Neurol. 2016;7:125.

18. Shah VP, Oliveira J E Silva L, Farah W, Seisa MO, Balla AK, Christensen A, Farah M, Hasan B, Bellolio F, Murad MH. Diagnostic accuracy of the physical examination in emergency department patients with acute vertigo or dizziness: A systematic review and meta-analysis for GRACE-3. Acad Emerg Med. 2023;30(5):552–578.

19. Gerlier C, Hoarau M, Fels A, Vitaux H, Mousset C, Farhat W, Firmin M, Pouyet V, Paoli A, Chatellier G, Ganansia O. Differentiating central from peripheral causes of acute vertigo in an emergency setting with the HINTS, STANDING, and ABCD2 tests: A diagnostic cohort study. Acad Emerg Med. 2021 Dec;28(12):1368-1378. doi: 10.1111/acem.14337. Epub 2021 Jul 20. PMID: 34245635.

20. Vanni S, Nazerian P, Pecci R, Pepe G, Pavellini A, Casula C, de Curtis E, Ronchetti M, Vannucchi P, Bartolucci M. Timing for nystagmus evaluation by STANDING or HINTS in patients with vertigo/dizziness in the emergency department. Acad Emerg Med. 2023 May;30(5):592–594.

21. Newman-Toker DE, Kerber KA, Hsieh YH, et al. HINTS outperforms ABCD2 to screen for stroke in acute continuous vertigo and dizziness. Acad Emerg Med. 2013;20:986–996.

22. Tarnutzer AA, Edlow JA. Bedside Testing in Acute Vestibular Syndrome-Evaluating HINTS Plus and Beyond-A Critical Review. Audiol Res. 2023 Sep 1;13(5):670–685.

