## Supplementary material for "Diagnostic accuracy of the STANDING algorithm in patients with isolated vertigo/dizziness, a multicentre prospective study (STANDING-M)": Cover Letter

Dear Editor,

We wish to submit an original research article titled "Diagnostic Accuracy of the STANDING Algorithm in Patients with Isolated Vertigo/Dizziness: A Multicentre Prospective Study (STANDING-M)." This article is original and has not been previously published. It highlights the usefulness of STANDING, a clinical algorithm for the differential diagnosis of isolated vertigo, which has been validated in two monocentric studies conducted in Italy and France.

In this multicentre study, the STANDING algorithm accurately identified patients with isolated vertigo who require urgent evaluation for central diseases across various types of emergency departments. These findings underscore that the accurate classification of vertigo and dizziness in emergency settings relies predominantly on clinical evaluation, including the correct assessment of nystagmus.

Thank you for considering our submission.

Sincerely,

Dr. Mattia Ronchetti
