## Supplementary material for "Diagnostic accuracy of the STANDING algorithm in patients with isolated vertigo/dizziness, a multicentre prospective study (STANDING-M)": Study Protocol

### OBSERVATIONAL STUDY

| | |
|---|---|
| **Study title:** | Diagnostic accurancy of STANDING algorithm for differential diagnosis of vertigo in emergency department: a multicenter study. |
| **Protocol code:** | STANDING – M |
| **Protocol version:** | version 2.0 |
| **Date:** | 02.05.2022 |
| **Sponsor:** | Azienda USL Toscana Centro |
| **Coordinator center:** | Medicina d'Urgenza Empoli |
| **Main investigator:** | *Dr Simone Vanni*<br>*Direttore SOC Medicina d'Urgenza EMPOLI*<br>*Direttore Area della Formazione*<br>*Dipartimento Emergenza Urgenza e Area Critica AUTC.* |
| **Other investigators:** | Dr Simone Magazzini, Direttore Dipartimento Emergenza Urgenza e Area Critica AUTC,<br>Dr Maurizio Bartolucci, Direttore Dipartimento Diagnostica per immagini AUTC<br>Dr.ssa Paola Bartalucci, SOC Medicina d'Urgenza Empoli AUTC<br>Dr.ssa Claudia Casula, SOC Medicina d'Urgenza Empoli AUTC |

**Participating centers list**

| | |
|---|---|
| **Center name** | *Medicina D'Urgenza Prato, AUTC*<br>*Dott.ssa Ersilia De Curtis* |
| **Center name** | *Medicina d'Urgenza Viareggio, AUTNO*<br>*Dott Giuseppe Pepe* |
| **Center name** | *Medicina d'Urgenza, AOU-Careggi* |

*Dott Peiman Nazerian*

#### Contact information

| **Sponsor contact** | *Dr Simone Vanni*<br>** |
| --- | --- |

#### PROTOCOL APPROVAL

The investigators:

- approve this Protocol;
- declare that the study will be conducted in accordance with this protocol..

| | |
| --- | --- |
| Dott Simone Vanni | Date |
| Dott. Simone Magazzini | Date |
| Dott Maurizio Bartolucci | Date |

#### Index

#### Background

The term "vertigo" comes from the Latin word "vertere" (turning, spinning) and describes an illusory and unpleasant sensation of moving (1). Vertigo represents a common medical problem which afflicts about 20-30% of the population (2) and it is a frequent cause of abstention from work and disability (3). Moreover, it is the main problem for 1-3% of admittances to emergency department (4). In most cases it is provoked by a benign disease of inner ear (5,6), however it can be the main symptom of a more dangerous illness like ischemic or hemorrhagic stroke, cerebral neoplasm or demyelinating disease (7-9). According to many studies, the incidence of cerebrovascular disease in patients suffering from vertigo who come to emergency department is between 2 and 6% (10,11). Vertigo is the prevailing clinical problem in patients with misdiagnosed ischemic stroke (12) and a missed or belated diagnosis leads to an increase of mortality in the acute phase of disease. In the current state, two diagnostic algorithms have been proposed for the evaluation of acute vertigo, named with the acronyms HINTS (14-16) and STANDING (17). The former is characterized by high sensibility and specificity when utilized by a specialist physician (neuro otologist), but it seems to be cumbersome to use in emergency department. Conversely, the latter has been validated in this setting with a prospective medical study (18,19) and includes some features of HINTS. However, in addition to examination do acute vestibular syndrome, it comprises the evaluation of benign paroxysmal positional vertigo (the most common type of vertigo in emergency department), and of upright position (20). Therefore, it allows a correct interpretation of patients with balance disorder without nystagmus. Although in italian and international emergency departments the STANDING algorithm is used by some physicians after its validation (19) ), only a part of them employs this method in Tuscany. The proposed multicenter study will allow to evaluate its diagnostic accuracy and at the same time to compare the different use of resources and the incidence of adverse effects in two cohorts of patients.

#### Objectives of the study

Evaluating diagnostic accuracy and clinical usefulness of a simplified algorithm (STANDING) for differential diagnosis of acute vertigo in emergency department.

In particular, the aim is to verify:

1) As primary outcome measure: test sensitivity and specificity for the diagnosis of central vertigo

ed in particular ischemic stroke. Diffusion weighted imaging (DWI) magnetic resonance imaging is the reference standard.

2) As secondary outcomes :

a) Number of brain imaging tests (CT with and without contrast medium and MRI) and specialist consultations demanded exclusively during the patient's stay in the emergency department.

b) Number of adverse events (death from all causes, diagnosis of central vertigo, in particular ischemic stroke, need for revascularization interventions through systemic thrombolysis or transcatheter thrombectomy, neurosurgical intervention, re-entry in the emergency department for vertigo determining hospitalization) in the population during the follow-up of 1 month.

#### Study design

Observational study assessing diagnostic accuracy, no profit, based on two cohorts of patients (one group composed by patients evaluated with STANDING algorithm and the other group of patients evaluated with standard examination).
The start of the study is expected upon receipt of the necessary authorizations from the competent Ethics Committee. The study duration is one year (see statistical analysis).

#### Setting

The proposed study is conceived as multicenter and takes place in the emergency departments of Azienda USL Toscana Centro (AUTC) in the district of Empoli (as promoter center) and Prato, of Azienda USL Toscana Nord-Ovest (AUTNO) in the district of Viareggio, and in Emergency department of Azienda Ospedaliera Universitaria Careggi (AOUC).

##### Study population

The patients that are considered eligible for the study are those who are adult and presenting in emergency departments of centers involved in the study suffering from vertigo/disequilibrium.

The definition of vertigo is described by two English terms "vertigo" e "dizziness': according to the consensus of Barany Society (2009), they mean "the sensation of self-motion when no self-motion is occurring" and "the sensation of disturbed or impaired spatial orientation without a false or distorted sense of motion". Disequilibrium is considered as a perceived alteration of the maintenance of upright position.

Exclusion Criteria

1) patients unable to cooperate (who are affected by severe dementia or incapable to provide consensus)
2) patients affected by disease of cervical spine of trauma of this part of body that contraindicate the manipulation of neck, necessary for the assessment of nystagmus
3) impractical follow-up (1 month)
4) dying patient (less three estimated months to live)
5) patients under the age of 18 years
6) patients with neurologic deficit identified during triage examination (Cincinnati Prehospital stroke scale, CPSS>0) of suffering from another disease that can be the cause of dizziness/balance disorder (e.g. anemia, arrhythmia, hypoglycemia, alcoholic intoxication)
7) patients without symptoms at the time of examination
8) patients who deny the participation in the study.

#### Study outcomes

PRIMARY OUTOCOME

The sensitivity, specificity, negative and positive predictive values, negative and positive likelihood ratio (LR) of STANDING algorithm with their 95% confidence intervals are estimated in both cohort of patients subjected to algorithm e and control cohort, in order to assess the diagnostic accuracy of the method. Diffusion weighted imaging (DWI) magnetic resonance imaging is the reference standard except for patients with a contraindication to an MRI. In this case the reference standard test is a computed tomography (CT) with contrast medium and angiography study (CTA).

SECONDARY OUTCOME

a) Number of requests for diagnostic imaging test (CT or MRI) and specialist consultations during the stay in emergency department. The imaging tests done during the hospitalization or in any case after the emergency department phase within one month from the enrollment are excluded from the analysis.

b) Number of adverse events (death from all causes, diagnosis of central vertigo, in particular ischemic stroke, need for revascularization interventions through systemic thrombolysis or transcatheter thrombectomy, neurosurgical intervention, re-entry in the emergency department for vertigo determining hospitalization) in the population during the follow-up of 1 month.

#### Variables

INDEX TEST: STANDING

The test is a diagnostic algorithm composed by four items 1) evaluation the presence of spontaneous nystagmus 2) evaluation of trajectory and direction of nystagmus 3) evaluation of head impulse test (HIT) 4) evaluation of upright position **(S**pon**TAN**eous**, D**irection, h**i**t, standi**NG**: STANDING) (17-20)

1) evaluation the presence of spontaneous nystagmus (a constant nystagmus in a state of rest): this phase is essential to recognize the presence of an acute vestibular syndrome, characterized by vertigo, nausea, vomiting, postural instability and spontaneous nystagmus. The presence of nystagmus is evaluated in the main gaze directions with the patient supine for at least 5 minutes, with and without Frenzel glasses. If nystagmus is not present, provoking maneuvers are performed for the horizontal (Pagnini-McCure) and vertical (Dix-Hallpike) canals, both on the right and on the left. In case of absence of both spontaneous and positional nystagmus, phase 4 must still be evaluated.
2) If a spontaneous nystagmus is present, it is necessary to evaluate its direction: if the nystagmus is multidirectional (gaze-evoked type: it beats to the right with the right lateral gaze and beats to the left with the left lateral gaze) or vertical, it is indicative of central vertigo. If it is unidirectional, step 3 should follow.
3) When a vestibular sensory organ is damaged, the afferents from the unaffected side are not opposed. Therefore, when the head is rotated rapidly towards the affected side the eyes will initially be pushed towards the affected side and it will bring the gaze back to the desired aim immediately after a rapid corrective (saccadic) movement in opposite direction. When the saccadic movement is present, the HIT test is considered positive and indicates a

peripheral pathology. Whereas if the saccadic movement is not present, the test is considered negative and indicative of a central pathology (16). Phase 3 will be performed in all cases of unidirectional nystagmus.

4) In any case, the ability to maintain an upright position or to walk without assistance will be assessed. If it is impossible (Carmona Score 2/3) (20), the test will be indicative of central vestibular disease.

The index test (STANDING) is performed before the imaging exams. The results of the test will remain unknown to those who evaluate imaging exams, especially to those who will view the diffusion-weighted MRI performed within 1 month of admission to emergency department. The commission that evaluate the final diagnosis will be aware of all the tests performed by the patient including the STANDING in group A, neuroimaging exams and the final follow-up examination .

Training

The Test will be applied by trained emergency physicians. The training includes 5 hours of theoretical lessons with powerpoint presentation, viewing of explanatory videos and 3 hours of STANDING application on healthy volunteers. Furthermore, before enrolling patients in the study, the physicians must have already evaluated at least 10 patients under the supervision of an experienced physician (SV, PN, GP, EoC).

REFERENCE STANDARD

The reference standard is the diffusion weighted imaging (DWI) MRI, except for patients who have contraindications to the exam MRI and in this case the reference standard is computed tomography (CT) with contrast medium and angiographic study (CTA). The diagnosis of central vertigo in based on the presence of an acute brain injury in the posterior cerebral fossa detected either on neuroimaging exam performed in the emergency department or during the one-month follow-up. The presence of acute ischemic stroke is diagnosed in the presence of a restricted diffusion on MRI or an evident hypodense area with the characteristics of ischemic damage on CT, in a location consistent with the patient's symptoms. In case that it has not been possible to perform a brain MRI, the final diagnosis of central vertigo is defined by a panel of experts composed by an emergency physician (SV), a radiologist (MB) and a neurologist (PB) according to clinical and instrumental data, also considering any events that occurred during the follow-up and the one month examination. If a patient dies before having a neuroimaging exam, he is considered having a central vertigo.

#### Bias

In order to reduce information bias concerning the different definition of exposure and/or outcome in the two groups, the result of the diagnostic algorithm will remain unknown to the team that will define the final diagnosis of central vertigo and to the radiologist who will evaluate the brain MRI. To decrease selection bias, patients will be assigned by the triage nurse to physicians who apply STANDING or not equally (1: 1 ratio).

#### Study sample size

According to the data of prospective study of STANDING algorithm (18), the estimated prevalence of the central vertigo is about 10% of the sample. In order to include at least 20 patients with central vertigo per group and estimate the values of 95% confidence interval of the sensitivity within 10%, considering a follow-up drop-out of about 10%, we plan to enroll 220 patients per group. Considering that in one year the total number of visits in the 4 centers is about 200,000 patients, that patients with vertigo/disequilibrium represent about 1% of the accesses (2000), that about 10% of these have exclusion criteria according to points 1) to 5) (200 patients) (18), and that another 20% (400) is excluded for point 6) (non-vestibular cause), and also counting that another 20% of cases could be evaluated at a time when a trained physician is not present for the STANDING (400), it is expected to enroll 440 patients in about 6 months. Considering a different starting period of the study in every center with an increasing rate of enrollment, it is expected to conclude the enrollment 10 months after the start of the study and to conclude data collection at 12 months.

#### Enrollment

The inclusion/exclusion criteria will be evaluated by a triage nurse who will assign the patient to a physician who applies the STANDING protocol (STANDING cohort) or to a physician who does not apply it (control cohort).

The patients included will be managed by the referring physician of the emergency department regardless of participation in the study. Laboratory exams and image tests (CT of the head, MRI of the brain, Doppler study of the epiaortic vessels, etc.), consultations (audiological, neurological ...), therapy, hospitalization, observation or discharge of the patient from the emergency department will be at the discretion of the referring physician whether the STANDING protocol is applied or not. All included patients will be re-evaluated at 1 month by the study team to establish the final diagnosis (see REFERENCE STANDARDS). Patients discharged from the emergency department will be instructed to contact the hospital once the symptoms change (worsen and/or reappear).

#### Follow-up

All enrolled patients are re-evaluated one month ± one week after the index visit in emergency department. If it is possible, a face-to-face visit is preferred. Otherwise, the patient is contacted by phone in order to establish: 1) the persistence of vertigo, 2) the occurrence of adverse events: death from all causes, diagnosis of central vertigo in particular ischemic stroke, the need for systemic thrombolysis or mechanical thrombectomy, neurosurgical intervention, re-entry into the emergency department due to vertigo determining hospitalization. If the patient is unreachable, any re-access to the emergency department and/or hospitalizations for the same reason with a diagnosis of stroke or brain disease will be searched through the computerized medical records available in the hospitals with particular attention 1) to any cerebral revascularization interventions through systemic thrombolysis or transcatheter thrombectomy or 2) to any neurosurgical intervention. If it is not possible to obtain any data in the 3 months following the index visit, not even by interrogating the computer systems, the patient is considered lost to follow-up.

#### Definition of conclusion

For an individual patient, the study is concluded after the one month follow-up visit. The study will generally be concluded at the end of follow-up of patients expected by the sample size analysis

#### Data analysis

The data required for the study are collected in a data sheet planned before the start of study (see annex 10a and 10b). The data are collected in a database protected by an access key (annex 11), anonymously by each center and then sent to the sponsor center (SOC Medicina d'Urgenza Empoli, AUTC). The responsibility for archiving, processing and storing global data lies with the SOC Medicina d'Urgenza of Empoli.

#### Statistical plan

Continuous variables are expressed as mean ± standard deviation (OS) and dichotomous variables as percentages with 95% confidence interval. We evaluate the diagnostic accuracy of the STANDING test for the diagnosis of central vertigo by calculating the sensitivity, the specificity and the positive and negative predictive value with the 95% confidence intervals. The difference in sensitivity and specificity of the STANDING test compared to the control group is also evaluated. For the assessment of secondary outcomes, the percentages of neuroimaging exams, neurological or audiological consultations requested in emergency department and adverse events combined are compared in the STANDING group and the control group. Comparisons between dichotomous variables expressed as percentages are performed by $\chi 2$, or by Fisher's exact test for values <5%. Comparisons between continuous variables are performed by Student's t-test or nonparametric tests for variables with non- normal distribution. A $p<0.05$ in the two-tailed test is considered significant. Calculations are performed with SPSS software (version 17.0, SPSS, Chicago, Illinois, USA).
Missing data are reported and are not considered for statistical purposes.
Patients lost to follow-up are excluded from the statistical analysis.

#### Administrative aspects

##### Study financing

The study is promoted by the Azienda USL Toscana Centro, a non-profit institution it is non funded. There are no additional costs for the participating health centers.

#### Ethical considerations

The Sponsor undertakes to ensure that the study is conducted in compliance with the dictates of the Declaration of Helsinki, in accordance with this protocol and with the Good Clinical Practice (GCP). The Sponsor also undertakes to protect personal data, clinical or not, of the subjects involved in the study according to what is established by national legislation [Legislative Decree 196/2003] and by the provisions on the protection of personal data sanctioned by the European Regulation n. 2016/679 (GDPR, General Data Protection Regulation / General Data Protection Regulation).

**Acquisition of informed consent and data processing**

It is the responsibility of the investigators, or of subjects appointed by them, to obtain the informed consent of patients after adequate information about the purposes, methods, expected benefits and foreseeable risks of the study. The investigators must also inform the participants that the non-participation or the interruption will not cause prejudice or harm towards them.

The Sponsor undertakes to protect the clinical and non-clinical, personal data of the subjects involved in the study according to what is established by national legislation [Legislative Decree no. Lvo. 196/2003] and by the European Regulation n. 2016/679 (GDPR, General Data

Protection Regulation/General Regulations for Personal Data Protection). The collection and storage of biological material is not foreseen.

**Conflict of interest**

The investigators and the sponsor declare the absence of any financial conflict of interest about the matter.

#### Responsibility and publication policies

##### Role of sponsor and investigators

The main investigator and investigators of enrollment centers participated in delineating protocol and study design. The collection, management, analysis and interpretation of data will be the responsibility of the investigators.

##### Data ownership

The ownership of data belongs to the sponsor center and it is shared with the principal investigators and with the investigators leading the enrollment centers.

**Publication policies**

At the end of the data collection expected one year after the start of study, the data are analyzed. At the end of the analysis, the results of study will be made available both through communications at conferences and through publications in scientific journals.

#### Bibliography

1. Bisdorff A, Von Brevern M, Lempert T, Newman-Toker DE. Classification of vestibular symptoms: towards an international classification of vestibular disorders. J Vestib Res. 2009;19(1-2):1-13.
2. Karatas M. Central vertigo and dizziness: epidemiology, differential diagnosis, and common causes. Neurologist. 2008 Nov;14(6):355–64.
3. Skøien AK, Wilhemsen K, Gjesdal S. Occupational disability caused by dizziness and vertigo: a register-based prospective study. Br J Gen Pract. 2008 Set;58(554):619–23.
4. Crespi V. Dizziness and vertigo: an epidemiological survey and patient management in the emergency room. Neurol. Sci. 2004 Mar;25 Suppl 1:S24–25
5. Madlon-Kay DJ. Evaluation and outcome of the dizzy patient. J Fam Pract. 1985 Ago;21(2):109–13.
6. Herr RD, Zun L, Mathews JJ. A directed approach to the dizzy patient. Ann Emerg Med. 1989 Giu;18(6):664–72.
7. Lee H, Sohn S-I, Cho Y-W, Lee S-R, Ahn B-H, Park B-R, et al. Cerebellar infarction presenting isolated vertigo: frequency and vascular topographical patterns. Neurology. 2006 Ott 10;67(7):1178–83.
8. Cappello M, di Blasi U, di Piazza L, Ducato G, Ferrara A, Franco S, et al. Dizziness and vertigo in a department of emergency medicine. Eur J Emerg Med. 1995 Dic;2(4):201–11.
9. Huang CY, Yu YL. Small cerebellar strokes may mimic labyrinthine lesions. J. Neurol. Neurosurg. Psychiatr. 1985 Mar;48(3):263–5
10. Lam JM, Siu WS, Lam TS, Cheung NK, Graham CA, Rainer TH. The epidemiology of patients with dizziness in an emergency department. Hong Kong J Emerg Med.2006;13:133-139
11. Kerber KA, Brown DL, Lisabeth LD, Smith MA, Morgenstern LB. Stroke among patients with dizziness, vertigo, and imbalance in the emergency department: a population-based study. Stroke. 2006 Ott;37(10):2484–7.
12. Savitz SI, Caplan LR, Edlow JA. Pitfalls in the diagnosis of cerebellar infarction. Acad Emerg Med. 2007 Gen;14(1):63–8.
13. Tarnutzer AA, Berkowitz AL, Robinson KA, Hsieh YH, Newman-Toker DE. Does my dizzy patient have a stroke? A systematic review of bedside diagnosis in acute vestibular syndrome. CMAJ 2011;183-E571-92.

14. Kattah JC, Talkad AV, Wang DZ, Hsieh YH, Newman-Toker DE. HINTS to diagnose stroke in the acute vestibular syndrome: three-step bedside oculomotor examination more sensitive than early MRI diffusion-weighted imaging. Stroke. 2009 Nov;40(11):3504-10. doi: 10.1161/STROKEAHA.109.551234. Epub 2009 Sep 17.PMID: 19762709
15. Newman-Toker DE, Kerber KA, Hsieh YH, Pula J, Omron R, Tehrani ASS, Mantokoudis G. Hanley DF, Zee DS, Kattah JC. HINTS Outperforms ABCD2 to screen for stroke in acute continuous vertigo and dizziness. Acad Emerg Med. 2013 Oct;20(10):986-96. doi: 10.1111/acem.12223.PMID: 24127701
16. Kattah JC . Use of HINTS in the acute vestibular syndrome. An Overview. Stroke Vasc Neurol. 2018 Jun 23;3(4):190-196. doi: 10.1136/svn-2018-000160. eCollection 2018 Dec.PMID: 30637123
17. Vanni S, Pecci R, Edlow JA, Nazerian P, Santimone R, Pepe G, Moretti M, Pavellini A, Caviglioli C, Casula C, Bigiarini S, Vannucchi P, Grifoni S. STANDING, a four-step bedside algorithm for differential diagnosis of acute vertigo in the Emergency Department. Fontiers Neurol 2017 Nov 7;8:590. doi: 10.3389/fneur.2017.00590. eCollection 2017.PMID: 29163350.
18. Vanni S, Pecci R, Casati C, Moroni F, Risso M, Ottaviani M, Nazerian P, Grifoni S, Vannucchi P. The STANDING, a bedside four-step algorithm for the differential diagnosis of acute vertigo in the Emergency Department. Acta Otorhinolaryngol Ital. 2014 Dec;34(6):419-26.
19. Gerlier C, Hoarau M, Fels A, Vitaux H, Mousset C, Farhat W, Firmin M, Pouyet V, Paoli A, Chatellier G, Ganansia O. Differentiating central from peripheral causes of acute vertigo in an emergency setting with the HINTS, STANDING, and ABCD2 tests: A diagnostic cohort study. Acad Emerg Med. 2021 Dec;28(12):1368-1378. doi: 10.1111/acem.14337. Epub 2021 Jul 20.PMID: 34245635
20. Carmona S, Martínez C, Zalazar G, Moro M, Batuecas-Caletrio A, Luis L, Gordon C. The Diagnostic Accuracy of Truncal Ataxia and HINTS as Cardinal Signs for Acute Vestibular Syndrome. Front Neurol. 2016 Aug 8;7:125. doi: 10.3389/fneur.2016.00125. eCollection 2016.PMID: 27551274
